# Epigenome-wide scan identifies differentially methylated regions for lung cancer using pre-diagnostic peripheral blood

**DOI:** 10.1101/2021.02.04.21251138

**Authors:** Naisi Zhao, Mengyuan Ruan, Devin C. Koestler, Jiayun Lu, Carmen J. Marsit, Karl T. Kelsey, Elizabeth A. Platz, Dominique S. Michaud

## Abstract

**Background:** To reduce lung cancer burden in the US, a better understanding of biological mechanisms in early disease development could provide new opportunities for risk stratification.

**Methods:** In a nested case-control study, we measured blood leukocyte DNA methylation levels in pre-diagnostic samples collected from 430 men and women in the 1989 CLUE II cohort. Median time from blood drawn to diagnosis was 14 years for all participants. We compared DNA methylation levels by case/control status to identify novel genomic regions, both single CpG sites and differentially methylated regions (DMRs), while controlling for known DNA methylation changes associated with smoking using a previously described pack-years based smoking methylation score. Stratification analyses were conducted by time from blood draw to diagnosis, histology, and smoking status.

**Results:** We identified sixteen single CpG sites and forty DMRs significantly associated with lung cancer risk (q < 0.05). The identified genomic regions were associated with genes including *H19, HOXA4, RUNX3, BRICD5, PLXNB2*, and *RP13*. For the single CpG sites, the strongest association was noted for cg09736286 in the *DIABLO* gene (OR [for 1 SD] = 2.99, 95% CI: 1.95-4.59, P-value = 4.81 × 10^−7^). For the DMRs, we found that CpG sites in the *HOXA4* region were hypermethylated in cases compared to controls.

**Conclusion:** The single CpG sites and DMRs that we identified represented significant measurable differences in lung cancer risk, providing new insights into the biological processes of early lung cancer development and potential biomarkers for lung cancer risk stratification.

## Introduction

Despite substantial reductions in lung cancer incidence and death rates over the past three decades (Siegel et al. 2020), lung cancer continues to be the leading cause of cancer death in the US and is projected to account for 135,720 deaths in 2020, approximately 22% of all cancer deaths (Siegel et al. 2020). To reduce the lung cancer burden in the US, cancer prevention and early detection remain a top priority (Moyer and Force 2014). However, while the conventional lung cancer screening method of using low-dose CT (LDCT) scans is effective in reducing lung cancer mortality (National Lung Screening Trial Research et al. 2011), it leads to high false positives rates (>95% of pulmonary nodules detected are benign) (Fabrikant et al. 2018), over-diagnosis (Heleno et al. 2018; Patz et al. 2014), radiation exposure (McCunney and Li 2014), and has had poor uptake (Quaife et al. 2016). A better understanding of biological mechanisms in early disease development, especially understanding gained through using DNA methylation data, may contribute new insights into lung cancer development and provide new opportunities for risk stratification, screening prioritization, and drug development.

Genome-wide DNA methylation profiling has provided new insight into risk factors, biological pathways, and disease processes. For instance, differences in blood leukocyte DNA methylation levels (proportion of CpGs methylated) have been associated with smoking (Baglietto et al. 2017; Shenker et al. 2013), elevated subclinical inflammation (Ahsan et al. 2017; Ligthart et al. 2016), obesity (Wahl et al. 2016; Xu et al. 2018), type II diabetes (Chambers et al. 2015), and heart disease (Agha et al. 2019; Huan et al. 2019). With respect to lung cancer, many of the alterations in blood leukocyte DNA methylation levels identified to date have been directly linked to smoking behaviors (Baglietto et al. 2017; Fasanelli et al. 2015; Shenker et al. 2013; Zhang et al. 2016b; Zhang et al. 2016c). One recent meta-analysis that included over 15,000 individuals identified 2,623 differentially methylated CpG sites that are related to cigarette smoking (Joehanes et al. 2016). Some of the identified smoking-related CpG sites have also been shown to mediate the effects of smoking on lung cancer (Baglietto et al. 2017; Battram et al. 2019; Fasanelli et al. 2015). Nevertheless, very few studies have investigated methylation changes associated with lung cancer risk that are not associated with smoking (Zhang et al. 2016a; Zhang et al. 2016b).

While smoking remains the strongest risk factor for lung cancer, identifying DNA methylation alterations that are not caused by active smoking could reveal important biological pathways. Among smokers, variation in methylation may indicate different genetic susceptibility to lung cancer, since individuals often differ in their ability to detoxify carcinogenic compounds or to repair induced DNA damage, for example. Alternatively, variation in methylation could be a result of other environmental risk factors, such as occupational exposures, changes in the immune response, radon, or secondhand smoke. Using blood samples collected men and women without a cancer diagnosis in 1989 in the CLUE II cohort (Genkinger et al. 2004; Kakourou et al. 2015), we compared DNA methylation levels in individuals who later developed lung cancer with those who did not develop lung cancer in the same time frame. Specifically, we aimed to identify both single CpG sites, and differentially methylated regions, measured in pre-diagnostic blood samples of lung cancer cases and matched controls, that represent measurable differences in lung cancer risk independent of smoking exposures.

## Materials and Methods

### Study population – CLUE I/II cohort

Subjects for this study were selected from among participants of the CLUE II cohort who had also participated in the CLUE I cohort study (flowchart in Supplemental Figure 1 and additional details in Supplemental Methods). Both cohorts were based in Washington County, MD, and were initially established to identify serological precursors to cancer and other chronic diseases (Genkinger et al. 2004; Kakourou et al. 2015; Schober et al. 1987). CLUE II was conducted from May through October 1989, during which 32,894 individuals (25,076 were Washington County residents) provided a blood sample (Comstock et al. 1991). Among all participants, 98.3% were white, reflecting the population of this county at the time, and 59% were female. Participants provided health information at baseline, including the potential confounders attained education, cigarette smoking status, number of cigarettes smoked daily, cigar/pipe smoking status, and self-reported weight and height, from which body mass index (BMI) was calculated.

### Lung cancer ascertainment

All incident lung cancer cases (ICD9 162 and ICD10 C34) were ascertained from linkage to the Washington County cancer registry (before 1992 to the present) and the Maryland Cancer Registry (since 1992 when it began to the present). We selected all 241 first primary incident lung cancer cases who had participated in CLUE I and were diagnosed after the day of blood draw in CLUE II through January 2018. Using incidence density sampling, we selected 1 control per case matching on age, sex, cigarette smoking status, number of cigarettes smoked daily, cigar/pipe smoking status, and date of CLUE II blood draw.

### DNA methylation measurements

DNA extracted from buffy coat was bisulfite-treated using the EZ DNA Methylation Kit (Zymo) and DNA methylation was measured at specific CpG sites across the genome using the 850K Illumina Infinium MethylationEPIC BeadArray (Illumina, Inc, CA, USA) at the University of Minnesota Genomic Center (details in Supplemental Methods).

### Statistical analysis

In the current nested case-control study, we aimed to identify novel genomic regions, both single CpG sites and differentially methylated regions where differences in DNA methylation levels are not explained by smoking exposures, by controlling for the known DNA methylation changes associated with smoking in the statistical analysis. To examine the association between CpG-specific DNA methylation and lung cancer risk, we conducted epigenome-wide association analysis using unconditional multivariable logistic regression to estimate odds ratios (OR) of lung cancer per 1 SD increase in methylation level at single CpG sites. To maximize power, we used unconditional logistic regression to include cases and controls without a matched pair, and included participants every time they were sampled. All models were adjusted for age at blood draw, sex, surrogate variables for batch effects (Leek et al. 2012; Leek et al. 2010), smoking status (never, former, current), pack-years based smoking methylation score (details in Supplemental Methods), BMI, and leukocyte cell composition (Houseman et al. 2012; Salas et al. 2018) (given the potential for confounding by cell composition) (Adalsteinsson et al. 2012). All p-values were adjusted for multiple comparisons using the false discovery rate (FDR) method. Analyses of single CpGs with lung cancer were also stratified by smoking status and time from blood-draw to diagnosis, and separately by non-small cell (NSCLC) and small cell (SCLC) histology. All controls were included in these three types of stratification analyses. All statistical analyses were performed in R (version 3.5.0).

We used the DMRcate Bioconductor R package (Peters et al. 2015) to identify differentially methylated regions (DMRs) associated with lung cancer risk. Adjusting for the same covariates as in the single CpG analyses, DMRs were calculated using a parameter setting of lambda=1,000 and kernel adjustment C=2 (default setting) (Peters et al. 2015). Statistically significant DMRs were required to have a minimum of two statistically significant single CpGs and to meet the multiple testing adjustment criteria of FDR<0.1. Associations were also examined by time from blood-draw to diagnosis and lung cancer histology. Two of the most statistically significant regions were further evaluated for patterns by time to diagnosis.

## Results

### Population characteristics

Table 1 presents the characteristics of the 208 lung cancer cases and 222 controls that we included in this study. Over 99% of the majority of participants were white. The median time to lung cancer diagnosis was 14 years. The median age at blood draw in 1989 was 59 and 57 years in cases and controls, respectively. Overall, 55% of cases and controls were women and 11% were never smokers (Table 1).

**Table 1.** Pre-diagnostic characteristics of lung cancer cases and matched controls nested in the CLUE II cohort study.

|  | Lung Cancer Cases | Controls |
| --- | --- | --- |
| N | 208 | 222 |
| Median Age (Range, Years) | 59 (30-83) | 57 (28-81) |
| Sex |  |  |
| Male | 95 (46%)<sup>b</sup> | 98 (44%) |
| Female | 113 (54%) | 124 (56%) |
| Cigarette Smoking Status |  |  |
| Never Smoker | 22 (11%) | 22 (10%) |
| Ever Smoker | 80 (38%) | 88 (40%) |
| Current Smoker | 106 (51%) | 112 (50%) |
| Median Smoking Intensity (Range, Cig/Day)<sup>a</sup> | 20 (0-80) | 20 (0-80) |
| Cigar or Pipe Smoking |  |  |
| Never | 178 (86%) | 189 (85%) |
| Ever | 30 (14%) | 33 (15%) |
| Median BMI, kg/m<sup>2</sup> (Range) | 25.6 (15.5-42.6) | 25.9 (18.0-40.9) |
| Median Time before diagnosis (Range, Years) | 14 (0-29) | -- |

<sup>a</sup> 2 missing in cases and 2 missing in controls
<sup>b</sup> Percentages were calculated among cases or among controls

### Single CpG EWAS analysis

The EWAS analysis identified 16 differentially methylated CpGs that were statistically significant after multiple comparisons correction (q<0.05). Results are presented in Table 2 (statistically significant CpGs were sorted by q-value) and Figure 1. Among the 16 CpGs, many were located in genomic regions that have been previously associated with lung cancer or other malignancies (*RUNX3* (Sato et al. 2005), *H19* (Huang et al. 2018), *BAIAP2L2* (Liu et al. 2020), *GPR132* (Chen et al. 2017), *CUEDC1* (Lopes et al. 2018), *SSBP4* (Guo et al. 2018), *AMPD2* (Gao et al. 2020), *ADAM11* (Sieuwerts et al. 2005), and *RTN4R* (He et al. 2020); the top 1000 CpGs based on q-value are presented in Supplemental Table 1).

**Figure 1.**
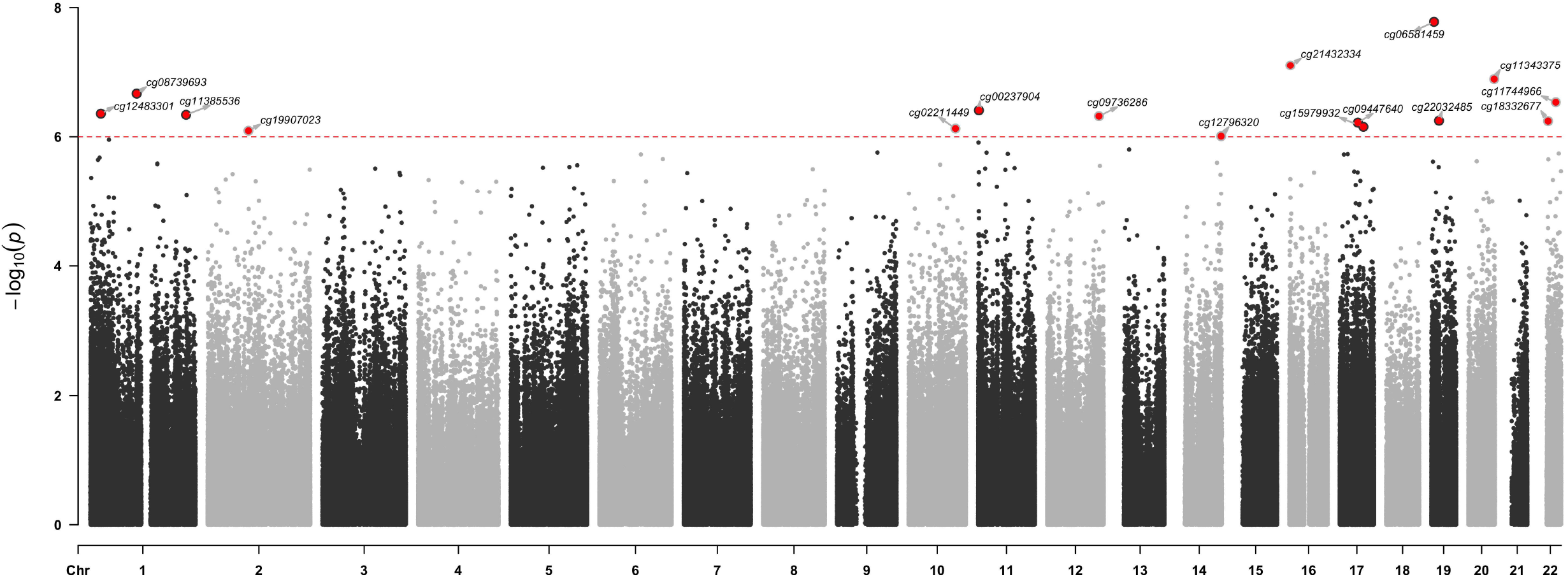
Manhatten plot for significant DNA-methylation (CpG) probes. CpGs reaching statistical significance are annotated (located above the dotted red horizontal line [p< 1 × 10−6]).

**Table 2.** Association between single CpGs and lung cancer, CLUE II: statistically significant CpGs after multiple comparisons correction (q<0.05)

| CpG | Annotation |  |  | Methylation Beta Value |  |  |  | Odds Ratio per 1 SD increase (95% CI)<sup>a</sup> |  |  |
| --- | --- | --- | --- | --- | --- | --- | --- | --- | --- | --- |
|  | Chr | Position | Gene | Mean in | Mean in | SD | Diff/SD | Overall | Time to diagnosis |  |
|  |  |  |  | Case | Control |  |  |  | ≤ 10 years | > 10 years |
| cg06581459 | chr19 | 6577668 |  | 0.606 | 0.585 | 0.055 | 0.383 | 2.26(1.70, 3.01) | 2.27(1.55, 3.32) | 2.15(1.54, 2.99) |
| cg21432334 | chr16 | 2262293 | BRICD5, PGP, C16orf79, RP11 | 0.819 | 0.809 | 0.034 | 0.312 | 2.28(1.69, 3.07) | 2.37(1.56, 3.60) | 2.10(1.48, 2.97) |
| cg11343375 | chr20 | 61268342 | RP13 | 0.673 | 0.657 | 0.047 | 0.343 | 2.42(1.74, 3.36) | 2.27(1.44, 3.59) | 2.44(1.66, 3.59) |
| cg08739693 | chr1 | 110165726 | AMPD2 | 0.785 | 0.776 | 0.036 | 0.256 | 2.17(1.62, 2.92) | 2.28(1.52, 3.41) | 2.21(1.56, 3.13) |
| cg11744966 | chr22 | 38506771 | BAIAP2L2 | 0.949 | 0.942 | 0.014 | 0.508 | 1.87(1.47, 2.37) | 2.04(1.45, 2.88) | 1.85(1.39, 2.46) |
| cg00237904 | chr11 | 2020314 | H19 | 0.519 | 0.489 | 0.076 | 0.401 | 1.94(1.50, 2.51) | 1.78(1.27, 2.49) | 1.87(1.40, 2.50) |
| cg12483301 | chr1 | 25239107 | RUNX3 | 0.697 | 0.678 | 0.058 | 0.333 | 2.81(1.88, 4.19) | 3.05(1.70, 5.47) | 2.72(1.72, 4.29) |
| cg11385536 | chr1 | 227133801 | CABC1, ADCK3 | 0.719 | 0.698 | 0.056 | 0.369 | 2.10(1.57, 2.80) | 2.00(1.34, 2.98) | 2.22(1.57, 3.12) |
| cg09736286 | chr12 | 122712925 | DIABLO | 0.867 | 0.849 | 0.056 | 0.33 | 2.99(1.95, 4.59) | 3.10(1.67, 5.74) | 3.33(1.97, 5.64) |
| cg22032485 | chr19 | 18531769 | SSBP4 | 0.632 | 0.615 | 0.062 | 0.269 | 2.47(1.73, 3.52) | 2.63(1.60, 4.30) | 2.23(1.49, 3.33) |
| cg18332677 | chr22 | 20239823 | RTN4R | 0.826 | 0.806 | 0.061 | 0.327 | 2.77(1.86, 4.13) | 2.34(1.35, 4.07) | 2.80(1.75, 4.48) |
| cg09447640 | chr17 | 42854853 | ADAM11 | 0.664 | 0.647 | 0.058 | 0.303 | 2.37(1.69, 3.34) | 3.44(2.05, 5.79) | 1.99(1.36, 2.92) |
| cg15979932 | chr17 | 55980107 | CUEDC1 | 0.812 | 0.796 | 0.04 | 0.389 | 2.01(1.53, 2.66) | 2.36(1.54, 3.62) | 1.84(1.34, 2.53) |
| cg02211449 | chr10 | 110671609 |  | 0.126 | 0.143 | 0.046 | -0.361 | 0.40(0.28, 0.58) | 0.46(0.28, 0.77) | 0.33(0.21, 0.51) |
| cg19907023 | chr2 | 96803532 | ASTL | 0.839 | 0.823 | 0.043 | 0.379 | 2.10(1.56, 2.81) | 1.76(1.18, 2.63) | 2.23(1.57, 3.15) |
| cg12796320 | chr14 | 105530592 | GPR132 | 0.504 | 0.487 | 0.059 | 0.279 | 2.20(1.60, 3.01) | 2.02(1.28, 3.18) | 2.41(1.65, 3.52) |

<sup>a</sup> Model adjusted for age at blood draw, sex, smoking status (never, former, current), pack-years based methylation score, BMI, surrogate variables for batch effects, and cell proportions.

CpGs previously reported to be associated with lung cancer risk, including cg05575921 in the *AHRR* gene (Fasanelli et al. 2015), cg03636183 in the *F2RL3* gene (Fasanelli et al. 2015), cg23387569 in the *AGAP2* gene (Baglietto et al. 2017), cg10151248 in the *PC* gene (Sandanger et al. 2018), and cg13482620 in the *B3GNTL1* gene (Sandanger et al. 2018), were not statistically significant in our analyses in which we adjusted for a pack-years methylation score. Adjusting for smoking status (never, former, current) but not for the packyears methylation score, two CpGs in the *AHRR* and *F2RL3* genes had similar-sized associations with lung cancer risk as previously reported (*AHRR*: OR [for 1 SD] = 0.43, 95% CI: 0.31-0.60, P-value = 6.76 × 10^−7^ vs. previously reported OR [for 1 SD] = 0.39, 95% CI: 0.24-0.61, P-value = 2.55 × 10^−5^ for cg05575921; *F2RL3*: OR for [1 SD] = 0.53, 95% CI: 0.40-0.70, P-value = 7.91 × 10^−6^ vs. previously reported OR [for 1 SD] = 0.51, 95% CI: 0.35-0.73, P-value = 4.19 × 10^−4^ for cg03636183) (Fasanelli et al. 2015). In addition, in the complete EWAS analysis conducted without adjusting for the packyears methylation score, only one CpG (cg14391737) was statistically significantly associated with lung cancer risk after adjusting for multiple comparisons. This CpG has been related to smoking in multiple studies (Joehanes et al. 2016) (top 1000 CpGs presented in Supplemental Table 2).

To further examine the associations of the significant CpGs with lung cancer risk, we stratified by time from blood draw to diagnosis (≤10, >10 years). The magnitude of risk was similar in the two strata; small differences in risk were likely due to statistical variability (Table 2). For these 16 differentially methylated CpGs, the ORs of lung cancer were in general slightly higher for former smokers and for SCLC, than among current smokers or for NSCLC, respectively (Supplemental Table 3).

### DMR analysis

Using the DMRcate package in R, we identified differentially methylated regions (DMRs) by case/control status (Peters et al. 2015). Instead of focusing on single CpG identification, the DMRcate method identifies regions of chromosomes that are differentially methylated by case-control status. After adjusting for both smoking status and packyears methylation score, forty DMRs were found to be statistically significantly associated with lung cancer risk (Table 3). Many of the top regions identified included genes that have been linked to lung cancer in previous studies (*H19* (Huang et al. 2018), *HOXA4* (Xu et al. 2019), *PLXNB2* (Liu and Zhao 2019), *PRDM1*(Zhu et al. 2017), *TSPAN4* (Ying et al. 2019), *PHPT1*(Xu et al. 2010), *MSI2* (Kudinov et al. 2016), *CBX5* (Yu et al. 2012), *RCAN1* (Ma et al. 2017), *CCL5* (Huang et al. 2009), and *BRDT* (Grunwald et al. 2006)).

**Table 3.** Association between differential methylated regions (DMR) and lung cancer in CLUE II participants ^a^.

| seqnames | start | end | width | no.cpgs | minfdr | maxbetafc | meanbetafc | overlapping.promoters |
| --- | --- | --- | --- | --- | --- | --- | --- | --- |
| chr1 | 6646079 | 6646156 | 78 | 3 | 5.15E-06 | 0.00930873 | 0.00646122 | ZBTB48-007, ZBTB48-002, ZBTB48-003 |
| chr1 | 28261367 | 28261706 | 340 | 8 | 2.45E-06 | -0.0078132 | -0.0039881 | SMPDL3B-002, SMPDL3B-001, SMPDL3B-003, SMPDL3B-004, SMPDL3B-006, SMPDL3B-007 |
| chr1 | 92414221 | 92414985 | 765 | 11 | 2.03E-10 | 0.03582019 | 0.0135806 | BRDT-008, BRDT-001, BRDT-002, BRDT-201, BRDT-202, BRDT-004, BRDT-005, BRDT-012, BRDT-003, BRDT-013, BRDT-007, BRDT-016, BRDT-017, BRDT-011, BRDT-009, BRDT-010, BRDT-015, BRDT-019, BRDT-020 |
| chr1 | 173174650 | 173174920 | 271 | 6 | 6.58E-06 | -0.0114329 | -0.005826 | TNFSF4-001, TNFSF4-002, TNFSF4-003 |
| chr1 | 206306094 | 206306498 | 405 | 4 | 3.37E-06 | -0.0156001 | -0.0114794 | RP11-38J22.6-001, RP11-38J22.6-003 |
| chr2 | 164204752 | 164205188 | 437 | 5 | 5.72E-06 | 0.02526139 | 0.0170268 | NA |
| chr3 | 6902824 | 6903067 | 244 | 6 | 7.67E-06 | -0.0183154 | -0.0088293 | GRM7-001, GRM7-004, GRM7-003, GRM7-201, GRM7-202, GRM7-007, GRM7-002, GRM7-013, GRM7-015 |
| chr3 | 71275993 | 71276631 | 639 | 5 | 2.44E-07 | 0.02400706 | 0.0153042 | NA |
| chr5 | 134783229 | 134783581 | 353 | 3 | 1.91E-06 | 0.00988655 | 0.00539191 | C5orf20-001 |
| chr6 | 27859476 | 27859811 | 336 | 3 | 2.27E-07 | 0.01242064 | 0.01074769 | HIST1H2BO-001, HIST1H3J-001, HIST1H2AM-001, HIST1H3J-002 |
| chr6 | 74364495 | 74364700 | 206 | 4 | 2.63E-06 | 0.02515135 | 0.02166436 | SLC17A5-001, SLC17A5-201 |
| chr6 | 106546074 | 106546824 | 751 | 8 | 4.62E-14 | 0.02624875 | 0.01650889 | PRDM1-002, PRDM1-007, PRDM1-003, RP1-134E15.3-001 |
| chr6 | 149805995 | 149806659 | 665 | 11 | 1.81E-08 | 0.01785178 | 0.01265741 | ZC3H12D-201, ZC3H12D-003, ZC3H12D-002, ZC3H12D-202, ZC3H12D-004, ZC3H12D-203 |
| chr7 | 2556962 | 2557715 | 754 | 10 | 1.14E-08 | 0.01226153 | 0.00494892 | LFNG-001, LFNG-201, LFNG-002, LFNG-004 |
| chr7 | 27169674 | 27171213 | 1540 | 20 | 3.16E-15 | 0.0371663 | 0.0205328 | HOXA4-001, HOXA4-201, HOXA4-004, HOXA-AS3-003 |
| chr7 | 110730805 | 110731201 | 397 | 3 | 3.21E-06 | -0.016941 | -0.014336 | LRRN3-003, LRRN3-001, LRRN3-002, LRRN3-201, LRRN3-004 |
| chr7 | 153584038 | 153584748 | 711 | 10 | 1.28E-07 | -0.020429 | -0.00886 | DPP6-001 |
| chr8 | 101225181 | 101225902 | 722 | 6 | 2.40E-07 | 0.01812174 | 0.01370006 | SPAG1-006 |
| chr9 | 136216873 | 136217303 | 431 | 5 | 1.35E-07 | 0.01245556 | 0.00840205 | RPL7A-005, RPL7A-009, MED22-002, MED22-006, SNORD36A-201, SNORD36B-201, MED22-003, SNORD24-201, RPL7A-004, MED22-005, MED22-010, RPL7A-001, RPL7A-003, MED22-004, MED22-009, RPL7A-006, RPL7A-007, RPL7A-008, MED22-011, RPL7A-002, SNORD36C-201 |
| chr9 | 139745116 | 139745412 | 297 | 5 | 5.79E-08 | 0.01178954 | 0.00625226 | PHPT1-001, MAMDC4-005, PHPT1-003, MAMDC4-201, PHPT1-005, PHPT1-004, MAMDC4-001, PHPT1-002, PHPT1-006, PHPT1-201 |
| chr10 | 63808314 | 63809170 | 857 | 14 | 4.11E-08 | 0.0110823 | 0.00745464 | ARID5B-002 |
| chr10 | 104535792 | 104536052 | 261 | 8 | 6.65E-06 | -0.0120183 | -0.0071156 | WBP1L-001 |
| chr11 | 847042 | 847072 | 31 | 2 | 7.57E-06 | 0.01410401 | 0.01263935 | TSPAN4-017, TSPAN4-005, TSPAN4-002, TSPAN4-012, TSPAN4-013, TSPAN4-018 |
| chr11 | 2019436 | 2021103 | 1668 | 31 | 1.22E-28 | 0.03449739 | 0.00903111 | H19-201, H19-008, H19-004, H19-002, H19-001, H19-005, H19-009, H19-012, H19-007, H19-013, H19-003, H19-006, H19-014 |
| chr11 | 128736844 | 128737467 | 624 | 8 | 1.17E-07 | -0.0292492 | -0.009666 | KCNJ1-002, KCNJ1-001, KCNJ1-005, KCNJ1-201, KCNJ1-003, KCNJ1-006 |
| chr12 | 54653403 | 54653788 | 386 | 4 | 6.51E-07 | -0.0179083 | -0.0145844 | CBX5-004, CBX5-003, CBX5-005 |
| chr16 | 2261947 | 2262293 | 347 | 6 | 5.27E-11 | 0.01398617 | 0.00891788 | BRICD5-001, RP11-304L19.8-001, PGP-003, BRICD5-002, BRICD5-003, BRICD5-004 |
| chr17 | 17464853 | 17465682 | 830 | 8 | 2.21E-09 | 0.02334441 | 0.01559593 | NA |
| chr17 | 34207332 | 34208012 | 681 | 6 | 4.60E-10 | 0.01767462 | 0.01343757 | CCL5-001, CCL5-002, CCL5-003 |
| chr17 | 55362520 | 55363072 | 553 | 4 | 5.30E-07 | -0.0150749 | -0.0124685 | MSI2-005 |
| chr19 | 17610751 | 17610785 | 35 | 2 | 2.91E-06 | -0.0122955 | -0.0107149 | SLC27A1-009, SLC27A1-010 |
| chr19 | 18209584 | 18209746 | 163 | 5 | 3.52E-06 | -0.0104941 | -0.0047016 | MAST3-001, IL12RB1-005, IL12RB1-006, IL12RB1-002 |
| chr20 | 36148133 | 36149750 | 1618 | 41 | 2.78E-17 | 0.02478338 | 0.0137516 | NNAT-001, NNAT-002, BLCAP-009, BLCAP-010, BLCAP-005 |
| chr20 | 62200050 | 62200711 | 662 | 6 | 6.94E-10 | 0.01497656 | 0.01241171 | HELZ2-001, HELZ2-006 |
| chr20 | 62688192 | 62688896 | 705 | 9 | 1.60E-06 | 0.01441642 | 0.01071442 | TCEA2-008, RP13-152O15.5-001 |
| chr21 | 35898717 | 35899468 | 752 | 8 | 5.29E-08 | 0.0200447 | 0.01593455 | RCAN1-003, RCAN1-201, RCAN1-005, RCAN1-007 |
| chr21 | 38362230 | 38363160 | 931 | 10 | 1.08E-08 | 0.04123433 | 0.01288671 | HLCS-001, HLCS-004 |
| chr22 | 31002362 | 31003283 | 922 | 14 | 1.55E-09 | -0.0113028 | -0.0050389 | TCN2-001, PES1-003, PES1-002, TCN2-002, TCN2-003, TCN2-004, TCN2-005, PES1-012, PES1-007 |
| chr22 | 50738160 | 50738890 | 731 | 3 | 2.49E-08 | 0.02463834 | 0.02309907 | PLXNB2-012 |
| chr22 | 51176117 | 51176734 | 618 | 7 | 1.57E-07 | 0.01094617 | 0.00757197 | ACR-001, AC000036.4-001, ACR-003, ACR-002 |

<sup>a</sup> Model adjusted for age at blood draw, sex, smoking status (never, former, current), pack-years based methylation score, BMI, surrogate variables for batch effects, and cell proportions.

We conducted stratified analyses to examine whether our DMR results were modified by time to diagnosis or differed by histology. Among those with ≤10 years between blood draw and diagnosis, a region located on chromosome 20:36148699-36149271 (genes *NNAT* and *BLCAP*) was statistically significantly associated with lung cancer. By histology, one region located in *H19* gene and one region located in *MYEOV* gene were statistically significant DMRs for NSCLC and SCLC, respectively (Supplemental Table 4).

We conducted further analyses for two of the most statistically significant DMRs. These two regions, located on the chromosome 11 (*H19* gene) and 7 (*HOXA4* gene), consisted of 31 and 20 CpGs, respectively, that differed between the cases and controls (all were q-value<0.1 FDR adjusted). For each of these two regions, we selected the CpGs with the strongest associations with lung cancer risk for comparison by time to cancer diagnosis (≤10, >10 years). Results for CpGs in these two regions are presented in Table 4 (top 10 statistically significant CpGs are included), with additional tables presented in Supplemental Table 5. For CpGs located in the *HOXA4* region, 18 out of 20 were statistically significant and all sites were hypermethylated in cases compared to controls. Results for these *HOXA4* region CpGs were similar in the ≤10 and >10 year time to diagnosis groups. In the *H19* region, the strongest association was noted for cg00237904. This CpG was also identified in the top statistically significant CpGs in the single CpG EWAS (Table 2, q-value<0.05).

**Table 4.**
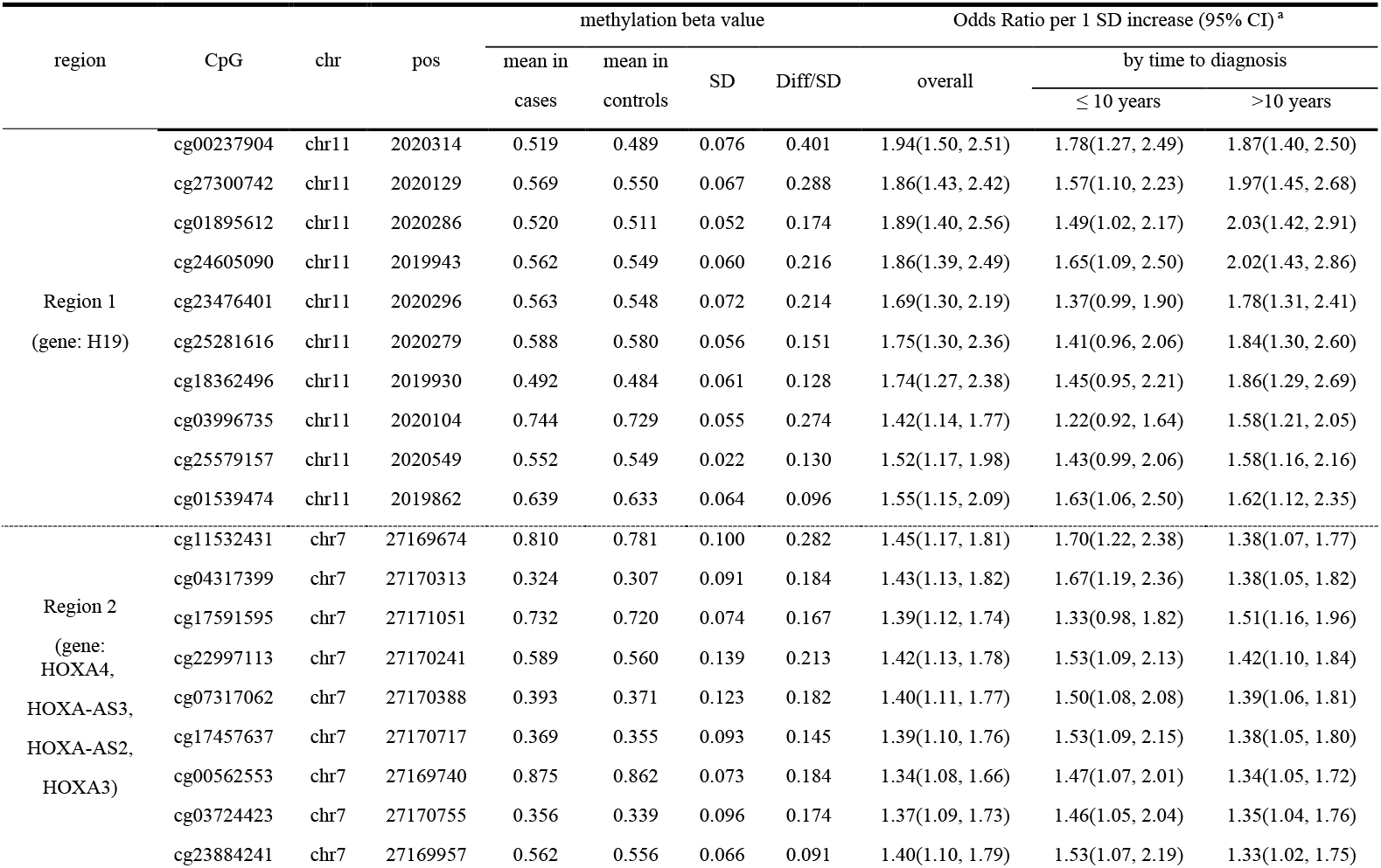
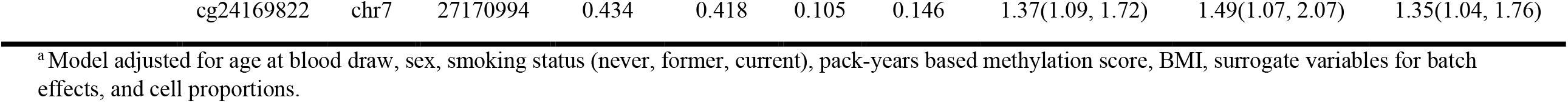
Odds ratio for top 10 CpGs (q-value<0.05) in two differentially methylated region (H19 and HOXA4) and lung cancer risk, and stratified by time to diagnosis in CLUE II participants.

## Discussion

In this study, we identified both single CpGs and DMRs in lung cancer that are not primarily driven by smoking history, by using a DNA methylation-based packyears score to adjust for cumulative smoking. Using a prospective study design (with pre-diagnostic blood), we identified 16 single CpG sites and 40 DMRs regions that were associated with lung cancer risk; genes in these regions included *H19, HOXA4, RUNX3, BRICD5, RP13*, and *PLXNB2*.

Previous studies have used either case-control or nested case-control designs to study the association between methylation markers and lung cancer risk. Retrospective case-control studies on this topic are not comparable to our study because they either used a different methodology to measure blood leukocyte methylation (Wang et al. 2010), or measured methylation biomarkers in sputum as a classifier for lung cancer risk (Leng et al. 2017; Liu et al. 2017). Three nested case-control studies previously used pre-diagnostic, peripheral blood samples to examine DNA methylation levels associated with lung cancer risk, while adjusting for smoking using self-reported information (Baglietto et al. 2017; Fasanelli et al. 2015; Sandanger et al. 2018). Fasanelli 2015 (Fasanelli et al. 2015) and Baglietto 2016 (Baglietto et al. 2017) together identified six CpGs (cg05951221, cg21566642, cg05575921, cg06126421, cg23387569 and cg03636183) with significant ORs for lung cancer after adjusting for smoking using self-reported smoking status. Further stratification showed that five of the six CpGs had methylation levels strongly influenced by smoking. Sandanger 2018 found two additional CpGs, cg10151248 (*PC*) and cg13482620 (*B3GNTL1*), to be significantly associated with lung cancer risk after adjusting for smoking status, pack-years, and a comprehensive smoking index built using self-reported information (Sandanger et al. 2018). In our analyses, cg05575921 (*AHRR*), cg03636183 (*F2RL3*), and cg21566642 methylation levels were statistically significantly associated with lung cancer when we adjusted for self-reported smoking status, whereas cg10151248 (*PC*), cg13482620 (*B3GNTL1*), and cg23387569 (*AGAP2*) methylation levels did not significantly differ between the cases and controls regardless of whether we adjusted for the pack-years methylation score or not. cg05951221 and cg06126421 were not associated with lung cancer.

Our analyses identified novel genomic regions that are independent of smoking exposures. Many of the significant single CpGs we identified through EWAS were located in genomic regions that have been previously associated with lung cancer or other malignancies (*RUNX3* (Sato et al. 2005), *H19* (Huang et al. 2018), *BAIAP2L2* (Liu et al. 2020), *GPR132* (Chen et al. 2017), *CUEDC1* (Lopes et al. 2018), *SSBP4* (Guo et al. 2018), *AMPD2* (Gao et al. 2020), *ADAM11* (Sieuwerts et al. 2005), and *RTN4R* (He et al. 2020)). For instance, the *RUNX3* region is a tumor suppressor gene that is implicated in lung cancer oncogenesis (Sato et al. 2005). Promoter hypermethylation of *RUNX3* has been associated with NSCLC survival (Yanagawa et al. 2007). Many of the top differentially methylated regions we identified using the DMRcate analysis included genes that have been previously linked to lung cancer in other studies (*H19* (Huang et al. 2018), *HOXA4* (Xu et al. 2019), *PLXNB2* (Liu and Zhao 2019), *PRDM1*(Zhu et al. 2017), *TSPAN4* (Ying et al. 2019), *PHPT1*(Xu et al. 2010), *MSI2* (Kudinov et al. 2016), *CBX5* (Yu et al. 2012), *RCAN1* (Ma et al. 2017), *CCL5* (Huang et al. 2009), and *BRDT* (Grunwald et al. 2006)), providing support for our findings. Of these eleven genetic regions already linked to lung cancer, many are shown to be connected to poor outcome in lung cancer. For instance, decreased expressed levels of *PLXNB2* (Liu and Zhao 2019) and *PRDM1* (Zhu et al. 2017) have been found to be correlated with poor prognosis in lung cancer, while *TSPAN4* (Ying et al. 2019), *PHPT1*(Xu et al. 2010), *MSI2* (Kudinov et al. 2016), and *CBX5* (Yu et al. 2012) are linked to metastasis. In addition, *HOXA4* (Xu et al. 2019), *RCAN1* (Ma et al. 2017), and *CCL5* (Huang et al. 2009) are involved in the growth, development, and migration of lung cancer cells. Other top regions identified in our study include genes that have been linked to breast cancer (*NNAT* (Nass et al. 2017), *RPL7A* (Zhu et al. 2001), and *HIST1H2BO* (Xie et al. 2019)), colorectal cancer (*RP11* (Sun et al. 2017; Wu et al. 2019)), endometrial cancer (*HELZ2* (Qiao et al. 2019)), pancreatic cancer (*LFNG* (Liu et al. 2016)), prostate cancer (*MAST3* (Dahlman et al. 2012)), renal cell carcinoma (*KCNJ1* (Guo et al. 2015)), and tumor progression (*ZC3H12D* (Huang et al. 2012)).

Of all the novel genomic regions that we identified as associated with lung cancer risk, the *H19* region was the only one that appeared in both of the single CpG EWAS and DMRcate results. The H19 long noncoding RNA (LncRNA) has been previously implicated in lung cancer causation. Inhibition of LncRNA H19 has been found to suppress the growth, migration, and invasion of NSCLC (Huang et al. 2018). In terms of disease development, loss of imprinting of the *H19* gene has been connected to a genome-wide loss of methylation, and associated with the transformation from normal to NSCLC (Anisowicz et al. 2008; Kondo et al. 1995; Langevin et al. 2015). In our analyses, we found that hypermethylation of many *H19* region CpGs were associated with lung cancer risk in CLUE II. The direction of this association was unexpected since the overexpression of H19 LncRNA in lung tumor is often correlated with hypomethylation of the promoter region CpGs (Kondo et al. 1995). H19 LncRNA belongs to a highly conserved imprinted gene cluster that plays important roles in embryonal development and growth control (Gabory et al. 2010) and *H19* region methylation has been found to be influenced by early life exposures, including maternal factors during pregnancy (Miyaso et al. 2017), suggesting the possibility that external exposures could impact *H19* methylation. Since the blood samples were drawn years before cancer diagnosis in this study, the methylation patterns we observed could be regions that are modulated early on in lung cancer development. More research is needed to investigate the methylation pattern in blood prior to cancer diagnosis.

Use of the DMRcate analysis methods and the Illumina Infinium MethylationEPIC 850K BeadArray allowed us to investigate genome-wide regional methylation level differences between lung cancer cases and controls. Some of the regions we identified that have not previously been associated with lung cancer should be investigated in other populations. It is possible that some of single CpGs and DMRs that we identified after adjusting for the packyear methylation score could be related to risk factors unique to CLUE II. Further studies are needed to investigate pathways related to the novel genomic regions that we identified.

This study demonstrated the importance of carefully controlling for known DNA methylation changes associated with smoking to be able to identify novel genomic regions. We showed the potential for this approach to identify DMRs (i.e., not single CpG alterations) by case/control status using peripheral blood collected prior to lung cancer diagnosis. These findings suggest that methylation changes detectable years prior to cancer diagnosis could potentially influence lung cancer risk, providing new insights into the biological processes of early lung cancer development. Further work in other populations should be conducted to validate regions that we observed to be associated with lung cancer risk independent of smoking exposures, especially among different ethnic and racial groups.

## Data Availability

The datasets generated during the current study are available from the corresponding author on reasonable request and will be deposited into dbGaP within 1 year.

## Acknowledgments

Cancer incidence data were provided by the Maryland Cancer Registry, Center for Cancer Surveillance and Control, Maryland Department of Health, 201 W. Preston Street, Room 400, Baltimore, MD 21201. We acknowledge the State of Maryland, the Maryland Cigarette Restitution Fund, and the National Program of Cancer Registries of the Centers for Disease Control and Prevention for the funds that helped support the availability of the cancer registry data.

## Declarations

### Authors’ contributions

DSM., KTK and EAP designed the study, obtained funding and acquisition of data. JL assisted with preparation of dataset. DSM supervised all research activities. MR and NZ conducted the statistical analyses. DSM and NZ drafted the manuscript. DSM., EAP, KTK, DCK and CJM interpreted the data and provided critical revisions of the manuscript. All authors read and approved the final version of the manuscript.

### Ethics approval

This study was approved by the Institutional Review Board at Johns Hopkins University Bloomberg School of Public Health and at Tufts University.

## References

Adalsteinsson BT et al. (2012) Heterogeneity in white blood cells has potential to confound DNA methylation measurements PLoS One 7:e46705 doi:10.1371/journal.pone.0046705

Agha G et al. (2019) Blood Leukocyte DNA Methylation Predicts Risk of Future Myocardial Infarction and Coronary Heart Disease Circulation 140:645–657 doi:10.1161/CIRCULATIONAHA.118.039357

Ahsan M et al. (2017) The relative contribution of DNA methylation and genetic variants on protein biomarkers for human diseases PLoS Genet 13:e1007005 doi:10.1371/journal.pgen.1007005

Anisowicz A et al. (2008) A high-throughput and sensitive method to measure global DNA methylation: application in lung cancer BMC cancer 8:222

Baglietto L et al. (2017) DNA methylation changes measured in pre-diagnostic peripheral blood samples are associated with smoking and lung cancer risk International journal of cancer Journal international du cancer 140:50–61 doi:10.1002/ijc.30431

Battram T et al. (2019) Appraising the causal relevance of DNA methylation for risk of lung cancer International journal of epidemiology 48:1493–1504

Chambers JC et al. (2015) Epigenome-wide association of DNA methylation markers in peripheral blood from Indian Asians and Europeans with incident type 2 diabetes: a nested case-control study The lancet Diabetes & endocrinology 3:526–534 doi:10.1016/S2213-8587(15)00127-8

Chen P et al. (2017) Gpr132 sensing of lactate mediates tumor–macrophage interplay to promote breast cancer metastasis Proceedings of the National Academy of Sciences 114:580–585

Comstock GW, Helzlsouer KJ, Bush TL (1991) Prediagnostic serum levels of carotenoids and vitamin E as related to subsequent cancer in Washington County, Maryland The American journal of clinical nutrition 53:260S–264S

Dahlman KB et al. (2012) Modulators of prostate cancer cell proliferation and viability identified by short-hairpin RNA library screening PLoS One 7:e34414

Fabrikant MS, Wisnivesky JP, Marron T, Taioli E, Veluswamy RR (2018) Benefits and Challenges of Lung Cancer Screening in Older Adults Clinical therapeutics doi:10.1016/j.clinthera.2018.03.003

Fasanelli F et al. (2015) Hypomethylation of smoking-related genes is associated with future lung cancer in four prospective cohorts Nature communications 6:1–9

Gabory A, Jammes H, Dandolo L (2010) The H19 locus: role of an imprinted non-coding RNA in growth and development Bioessays 32:473–480

Gao Q-Z, Qin Y, Wang W-J, Fei B-J, Han W-F, Jin J-Q, Gao X (2020) Overexpression of AMPD2 indicates poor prognosis in colorectal cancer patients via the Notch3 signaling pathway World Journal of Clinical Cases 8:3197

Genkinger JM, Platz EA, Hoffman SC, Comstock GW, Helzlsouer KJ (2004) Fruit, vegetable, and antioxidant intake and all-cause, cancer, and cardiovascular disease mortality in a community-dwelling population in Washington County, Maryland American journal of epidemiology 160:1223–1233 doi:10.1093/aje/

Grunwald C et al. (2006) Expression of multiple epigenetically regulated cancer/germline genes in nonsmall cell lung cancer International journal of cancer 118:2522–2528

Guo X et al. (2018) A comprehensive cis-eQTL analysis revealed target genes in breast cancer susceptibility loci identified in genome-wide association studies The American Journal of Human Genetics 102:890–903

Guo Z et al. (2015) KCNJ1 inhibits tumor proliferation and metastasis and is a prognostic factor in clear cell renal cell carcinoma Tumor Biology 36:1251–1259

He Y, Ji P, Li Y, Wang R, Ma H, Yuan H (2020) Genetic Variants Were Associated With the Prognosis of Head and Neck Squamous Carcinoma Frontiers in oncology 10:372

Heleno B, Siersma V, Brodersen J (2018) Estimation of Overdiagnosis of Lung Cancer in Low-Dose Computed Tomography Screening: A Secondary Analysis of the Danish Lung Cancer Screening Trial JAMA Intern Med 178:1420–1422 doi:10.1001/jamainternmed.2018.3056

Houseman EA et al. (2012) DNA methylation arrays as surrogate measures of cell mixture distribution BMC bioinformatics 13:86 doi:10.1186/1471-2105-13-86

Huan T et al. (2019) Genome-wide identification of DNA methylation QTLs in whole blood highlights pathways for cardiovascular disease Nat Commun 10:4267 doi:10.1038/s41467-019-12228-z

Huang C-Y, Fong Y-C, Lee C-Y, Chen M-Y, Tsai H-C, Hsu H-C, Tang C-H (2009) CCL5 increases lung cancer migration via PI3K, Akt and NF-κB pathways Biochemical pharmacology 77:794–803

Huang S et al. (2012) The putative tumor suppressor Zc3h12d modulates toll-like receptor signaling in macrophages Cellular signalling 24:569–576

Huang Z, Lei W, Hu HB, Zhang H, Zhu Y (2018) H19 promotes non-small-cell lung cancer (NSCLC) development through STAT3 signaling via sponging miR-17 Journal of cellular physiology 233:6768–6776

Joehanes R et al. (2016) Epigenetic signatures of cigarette smoking Circulation: cardiovascular genetics 9:436–447

Kakourou A et al. (2015) Interleukin-6 and risk of colorectal cancer: results from the CLUE II cohort and a meta-analysis of prospective studies Cancer Causes & Control 26:1449–1460

Kondo M, Suzuki H, Ueda R, Osada H, Takagi K, Takahashi T (1995) Frequent loss of imprinting of the H19 gene is often associated with its overexpression in human lung cancers Oncogene 10:1193–1198

Kudinov AE et al. (2016) Musashi-2 (MSI2) supports TGF-β signaling and inhibits claudins to promote non-small cell lung cancer (NSCLC) metastasis Proceedings of the National Academy of Sciences 113:6955–6960

Langevin SM, Kratzke RA, Kelsey KT (2015) Epigenetics of lung cancer Translational Research 165:74–90

Leek JT, Johnson WE, Parker HS, Jaffe AE, Storey JD (2012) The sva package for removing batch effects and other unwanted variation in high-throughput experiments Bioinformatics 28:882–883

Leek JT et al. (2010) Tackling the widespread and critical impact of batch effects in high-throughput data Nature Reviews Genetics 11:733–739

Leng S et al. (2017) Gene methylation biomarkers in sputum as a classifier for lung cancer risk Oncotarget 8:63978

Ligthart S et al. (2016) DNA methylation signatures of chronic low-grade inflammation are associated with complex diseases Genome Biol 17:255 doi:10.1186/s13059-016-1119-5

Liu D et al. (2017) The indirect efficacy comparison of DNA methylation in sputum for early screening and auxiliary detection of lung cancer: a meta-analysis International journal of environmental research and public health 14:679

Liu H, Zhao H (2019) Prognosis related miRNAs, DNA methylation, and epigenetic interactions in lung adenocarcinoma Neoplasma 66:487–493

Liu J, Shangguan Y, Sun J, Cong W, Xie Y (2020) BAIAP2L2 promotes the progression of gastric cancer via AKT/mTOR and Wnt3a/β-catenin signaling pathways Biomedicine & Pharmacotherapy 129:110414

Liu P et al. (2016) Quantitative secretomic analysis of pancreatic cancer cells in serum-containing conditioned medium Scientific reports 6:37606

Lopes R et al. (2018) CUEDC1 is a primary target of ERα essential for the growth of breast cancer cells Cancer letters 436:87–95

Ma N, Shen W, Pang H, Zhang N, Shi H, Wang J, Zhang H (2017) The effect of RCAN1 on the biological behaviors of small cell lung cancer Tumor Biology 39:1010428317700405

McCunney RJ, Li J (2014) Radiation risks in lung cancer screening programs Chest 145:618–624

Miyaso H, Sakurai K, Takase S, Eguchi A, Watanabe M, Fukuoka H, Mori C (2017) The methylation levels of the H19 differentially methylated region in human umbilical cords reflect newborn parameters and changes by maternal environmental factors during early pregnancy Environmental research 157:1–8

Moyer VA, Force USPST (2014) Screening for lung cancer: U.S. Preventive Services Task Force recommendation statement Ann Intern Med 160:330–338 doi:10.7326/M13-2771

Nass N et al. (2017) High neuronatin (NNAT) expression is associated with poor outcome in breast cancer Virchows Archiv 471:23–30

National Lung Screening Trial Research T et al. (2011) Reduced lung-cancer mortality with low-dose computed tomographic screening N Engl J Med 365:395–409 doi:10.1056/NEJMoa1102873

Patz EF, Jr. et al. (2014) Overdiagnosis in low-dose computed tomography screening for lung cancer JAMA Intern Med 174:269–274 doi:10.1001/jamainternmed.2013.12738

Peters TJ et al. (2015) De novo identification of differentially methylated regions in the human genome Epigenetics Chromatin 8:6 doi:10.1186/1756-8935-8-6

Qiao Z, Jiang Y, Wang L, Wang L, Jiang J, Zhang J (2019) Mutations in KIAA1109, CACNA1C, BSN, AKAP13, CELSR2, and HELZ2 are associated with the prognosis in endometrial cancer Frontiers in Genetics 10:909

Quaife SL et al. (2016) The Lung Screen Uptake Trial (LSUT): protocol for a randomised controlled demonstration lung cancer screening pilot testing a targeted invitation strategy for high risk and ‘hard-to-reach’patients BMC cancer 16:1–9

Salas LA, Koestler DC, Butler RA, Hansen HM, Wiencke JK, Kelsey KT, Christensen BC (2018) An optimized library for reference-based deconvolution of whole-blood biospecimens assayed using the Illumina HumanMethylationEPIC BeadArray Genome Biol 19:64 doi:10.1186/s13059-018-1448-7

Sandanger TM et al. (2018) DNA methylation and associated gene expression in blood prior to lung cancer diagnosis in the Norwegian Women and Cancer cohort Scientific reports 8:1–10

Sato K, Tomizawa Y, Iijima H, Saito R, Ishizuka T, Nakajima T, Mori M (2005) Epigenetic inactivation of the RUNX3 gene in lung cancer Oncology reports 15:129–135

Schober SE, Comstock GW, Helsing KJ, Salkeld RM, Morris JS, Rider AA, Brookmeyer R (1987) Serologic precursors of cancer: I. Prediagnostic serum nutrients and colon cancer risk American journal of epidemiology 126:1033–1041

Shenker NS et al. (2013) Epigenome-wide association study in the European Prospective Investigation into Cancer and Nutrition (EPIC-Turin) identifies novel genetic loci associated with smoking Hum Mol Genet 22:843–851 doi:10.1093/hmg/dds488

Siegel RL, Miller KD, Jemal A (2020) Cancer statistics, 2020 CA Cancer J Clin 70:7–30 doi:10.3322/caac.21590

Sieuwerts AM et al. (2005) How ADAM-9 and ADAM-11 differentially from estrogen receptor predict response to tamoxifen treatment in patients with recurrent breast cancer: a retrospective study Clinical Cancer Research 11:7311–7321

Sun L et al. (2017) Down-regulation of long non-coding RNA RP11-708H21. 4 is associated with poor prognosis for colorectal cancer and promotes tumorigenesis through regulating AKT/mTOR pathway Oncotarget 8:27929

Wahl S et al. (2016) Epigenome-wide association study of body mass index, and the adverse outcomes of adiposity Nature 541:81 doi:10.1038/nature20784 https://www.nature.com/articles/nature20784#supplementary-information

Wang L et al. (2010) Methylation markers for small cell lung cancer in peripheral blood leukocyte DNA Journal of Thoracic Oncology 5:778–785

Wu Y et al. (2019) m 6 A-induced lncRNA RP11 triggers the dissemination of colorectal cancer cells via upregulation of Zeb1 Molecular cancer 18:1–16

Xie W et al. (2019) Expression and potential prognostic value of histone family gene signature in breast cancer Experimental and therapeutic medicine 18:4893–4903

Xu A-j, Xia X-h, Du S-t, Gu J-c (2010) Clinical significance of PHPT1 protein expression in lung cancer Chinese medical journal 123:3247–3251

Xu K, Liu B, Ma Y (2019) The tumor suppressive roles of ARHGAP25 in lung cancer cells OncoTargets and therapy 12:6699

Xu K, Zhang X, Wang Z, Hu Y, Sinha R (2018) Epigenome-wide association analysis revealed that SOCS3 methylation influences the effect of cumulative stress on obesity Biol Psychol 131:63–71 doi:10.1016/j.biopsycho.2016.11.001

Yanagawa N, Tamura G, Oizumi H, Kanauchi N, Endoh M, Sadahiro M, Motoyama T (2007) Promoter hypermethylation of RASSF1A and RUNX3 genes as an independent prognostic prediction marker in surgically resected non-small cell lung cancers Lung cancer 58:131–138

Ying X, Zhu J, Zhang Y (2019) Circular RNA circ-TSPAN4 promotes lung adenocarcinoma metastasis by upregulating ZEB1 via sponging miR-665 Molecular genetics & genomic medicine 7:e991

Yu Y-H et al. (2012) Network biology of tumor stem-like cells identified a regulatory role of CBX5 in lung cancer Scientific reports 2:584

Zhang X, Gao L, Liu Z-P, Jia S, Chen L (2016a) Uncovering driver DNA methylation events in nonsmoking early stage lung adenocarcinoma BioMed research international 2016

Zhang Y, Breitling LP, Balavarca Y, Holleczek B, Schottker B, Brenner H (2016b) Comparison and combination of blood DNA methylation at smoking-associated genes and at lung cancer-related genes in prediction of lung cancer mortality Int J Cancer 139:2482–2492 doi:10.1002/ijc.30374

Zhang Y, Elgizouli M, Schottker B, Holleczek B, Nieters A, Brenner H (2016c) Smoking-associated DNA methylation markers predict lung cancer incidence Clin Epigenetics 8:127 doi:10.1186/s13148-016-0292-4

Zhu Y, Lin H, Li Z, Wang M, Luo J (2001) Modulation of expression of ribosomal protein L7a (rpL7a) by ethanol in human breast cancer cells Breast cancer research and treatment 69:29–38

Zhu Z, Wang H, Wei Y, Meng F, Liu Z, Zhang Z (2017) Downregulation of PRDM1 promotes cellular invasion and lung cancer metastasis Tumor Biology 39:1010428317695929

